# Cohort Profile: Baseline Characteristics of Veterans from Improving Veteran Access to Integrated Management of Back Pain (AIM-Back) – an Embedded Pragmatic, Cluster Randomized Trial in the United States

**DOI:** 10.1101/2024.11.23.24317833

**Authors:** Steven Z. George, Courtni France, Cynthia J. Coffman, Kelli D. Allen, Trevor A. Lentz, Rebecca North, Ashley Choate, Adam P. Goode, Corey B. Simon, Janet M. Grubber, Heather King, Chad E. Cook, Francis J. Keefe, Lindsay A. Ballengee, Jennifer Naylor, Joseph Leo Brothers, Catherine Stanwyck, Travis Linton, Christa Tumminello, S. Nicole Hastings

**Affiliations:** Duke Clinical Research Institute and Department of Orthopaedic Surgery, Duke University, Durham NC; Center of Innovation to Accelerate Discovery and Practice Transformation at Durham VAHCS, HSR Durham NC; Center of Innovation to Accelerate Discovery and Practice Transformation, Durham VAHCS, HSR; Department of Biostatistics and Bioinformatics, Duke University School of Medicine; Center of Innovation to Accelerate Discovery and Practice Transformation, Durham VAHCS, HSR, Durham NC; University of North Carolina-Chapel Hill, Chapel Hill NC; Center for the Study of Aging and Human Development, Duke University School of Medicine, Durham NC; Center of Innovation to Accelerate Discovery and Practice Transformation at Durham VAHCS, HSR, Durham NC; Duke Clinical Research Institute and Department of Orthopaedic Surgery, Durham NC; Duke Clinical Research Institute, Center for Aging and Department of Orthopaedic Surgery, Duke University, Durham NC; Center of Innovation to Accelerate Discovery and Practice Transformation at Durham VAHCS, HSR, Durham NC; Cooperative Studies Program Coordinating Center - VA Boston Health Care System; Center of Innovation to Accelerate Discovery and Practice Transformation at Durham VAHCS, HSR and Department of Population Health Sciences and Division of General Internal Medicine at Duke University, Durham NC; Department of Psychiatry and Behavioral Sciences, Duke University, Durham NC; Center of Innovation to Accelerate Discovery and Practice Transformation at Durham VAHCS, HSR; Department of Population Health Sciences, Duke University, Durham NC; Durham VAHCS, MIRECC and Duke University, Durham NC; Duke Clinical Research Institute, Durham NC; Department of Rehabilitation Services, Duke University, Durham NC; Center of Innovation to Accelerate Discovery and Practice Transformation at Durham VAHCS, HSR; Duke University School of Medicine, Division of Geriatrics; Department of Population Health Sciences, Duke University School of Medicine, Durham NC

## Abstract

**Purpose:** AIM-Back is an embedded pragmatic clinical trial (ePCT) with cluster randomization designed to increase access and compare the effectiveness of two different non-pharmacological care pathways for low back pain (LBP) delivered within the Veteran Administration Health Care System (VAHCS). This manuscript describes baseline characteristics of AIM-Back participants as well as the representativeness of those referred to the AIM-Back program by sex, age, race, and ethnicity, relative to Veterans with low back pain at participating clinics.

**Participants:** To be eligible for AIM-Back, Veterans were referred to the randomized pathway at their clinic by trained primary care providers (Referral cohort). Veterans from the Referral cohort that participated in the study included: 1) an Electronic Health Record (EHR) sample of Veterans enrolled in the program (i.e., attended initial AIM-Back visit with no consent required) and a Survey sample of Veterans that were consented for further study. Descriptive statistics for age, race, ethnicity, sex, high-impact chronic pain (HICP), a comorbidity measure, post-traumatic stress diagnosis (PTSD) and opioid exposure were reported for the Referral cohort and by sample; mean baseline PROMIS pain interference, physical function and sleep disturbance scores were reported by sample. Additional measures of pain, mental health and social risk were reported on the Survey sample. Participation to prevalence ratios (PPRs) were calculated for sex, age, race, and ethnicity by clinic to describe representativeness of the Referral cohort.

**Findings to Date:** Across 17 randomized primary care clinics, the Referral cohort included 2767 unique Veterans with n=1817 in the EHR sample, n=996 in the Survey sample and n=799 of the EHR sample (44%) were also in the Survey sample. High rates of HICP were observed in the EHR and Survey samples (>59%). Mean scores (SD) based on self-reported PROMIS Pain Interference (63.2 (6.8), 63.1 (6.6)) and PROMIS Physical Function (37.1 (5.3), 38.1 (5.8)) indicated moderate impairment in the EHR sample and Survey sample respectively. Approximately 10% of the EHR sample had documented opioid use in the year leading up to the AIM-Back referral. At most clinics, older Veterans (>=65 years) were underrepresented in the Referral cohort compared to those with LBP visits at clinics (PPRs < 0.8).

**Future Plans:** The AIM-Back trial will conduct analysis to examine the comparative effectiveness of the two care pathways and identify individual characteristics that may improve responses to each pathway. The trial is expected to complete 12-month follow-up data collection by December 2024, with subsequent analyses and publications providing insights into optimizing non-pharmacological care for Veterans with LBP.

**Trial Registration:** NCT04411420 (clinicaltrials.gov)

**Strengths and Limitations:** *Strengths:*

- Embedded features (e.g., EHR documentation and clinical staff serving as interventionist) of the AIM-Back trial have potential to facilitate implementation of these care pathways more broadly in Veteran’s Health Administration clinics and in health systems outside the Veteran’s Health Administration
- Participating clinics were in 10 states providing regional geographic representation across the study.
- Trial was delivered within an integrated health system optimal for consistent documentation templates and data extraction across the trial sites.

*Limitations:*

- Due to clinical personnel’s role in participant recruitment, sample biases may exist in those referred to the program and/or those who agreed to participate in additional surveys
- These findings may not be generalizable to older aged Veterans due to our lower enrollment of those 65+.

## Introduction

Non-pharmacological treatments have been endorsed as effective, low risk (i.e., compared to pharmacological or interventional) approaches for treating low back pain (LBP) by multiple entities inside and outside the U.S., including the Center for Disease Control and Prevention [1], National Academy of Medicine [2], American College of Physicians [3], and World Health Organization.[4] Furthermore, there is evidence from observational studies indicating that non-pharmacological treatments for LBP reduce risk of future opioid use.[5–7] Consensus recommendations in practice guidelines, and encouraging findings from the previously cited observational studies, provide foundational support for non-pharmacological care of LBP, but there is still an urgent need for research designs that address their effectiveness. Pressing research questions about the structuring of non-pharmacologic care pathways to optimize clinical outcomes such as pain interference and physical function persist.[8] Many of these research questions can only be addressed through pragmatic trials designed to assess the impact of non-pharmacological care delivered in real-world settings.[9,10]

Accordingly, we conducted the Improving Veteran Access to Integrated Management of Back Pain (AIM-Back) trial.[11] AIM-Back was designed as an embedded pragmatic clinical trial (ePCT) with cluster randomization and is part of the National Institutes of Health-Department of Defense-Veteran Affairs Pain Management Collaboratory. This Collaboratory focuses on active military and Veterans for clinical trials because of the higher severity from LBP or joint pain reported in these populations.[12] Specific to the purposes of AIM-Back, the widespread prevalence of LBP and the disproportionate impact it has on Veterans’ quality of life [13] indicated a need to develop and test the effectiveness of different non-pharmacological care pathways in the Veteran Health Administration.

The AIM-Back trial: 1) developed and implemented two different LBP care pathways designed to improve access to non-pharmacological treatments; 2) is investigating the comparative effectiveness of these care pathways for reducing pain interference and improving physical function; and 3) plans to identify individual level characteristics that improve outcome response to a care pathway (i.e. heterogeneity of treatment effect analyses). The purpose of this Cohort Profile is to provide an overview of trial enrollment and a description of Veteran characteristics of the AIM-Back trial at baseline.

Specifically, this manuscript includes a description of: a) baseline characteristics including rates of HICP and opioid use (which are planned subgroups for future analyses), b) baseline measures of pain and function, and c) the representativeness of those referred to AIM-Back by sex, age, race, and ethnicity, relative to the clinical LBP population at randomized clinics, to provide an indication of how generalizable findings from this trial will be. Veterans that participated in the study include: 1) an Electronic Health Record (EHR) sample that enrolled in a clinical program (no consent required) and 2) a Survey sample that agreed to be contacted and consented for further study. There are Veterans that are in both the EHR and Survey Samples as they were subsets of Veterans referred by primary care providers to AIM-Back. Collectively, the information presented in this paper will provide an in-depth summary of the cohort’s individual level characteristics and serve as important source material for interpretation of the AIM-Back trial’s primary and secondary findings.

## Cohort Description

### Study Design

AIM-Back (registered at clinicaltrials.gov: NCT04411420) is a multisite, ePCT that compares the effectiveness of two clinical care pathways for Veterans with LBP within the Veterans Health Administration (VHA) Health Care System. The AIM-Back trial has been described previously in our protocol paper, including a description of its pragmatic components.[11] Both pathways were initiated by a referral from a trained primary care provider and are briefly summarized below. The Sequenced Care Pathway (SCP) started with an evaluation and treatment by a local VA physical therapist. Veterans were then offered six sessions of weekly telehealth physical activity calls through a remote centralized care provider. Veterans then returned for a follow-up assessment with their local physical therapist, which included risk screening for persistent disability using the STarT Back Screening Tool (SBST).[14] Those identified as medium or high risk on the SBST received an additional six weeks of remote care focused on pain coping skills training delivered by physical therapists trained in psychologically informed care. Additional details for the SCP and metrics for its usage have been previously described.[15]

The Pain Navigator Pathway (PNP) involved referral of the Veteran to a Pain Navigator, i.e., a local VA provider that worked with Veterans primarily via telehealth to identify appropriate non-pharmacologic treatment options, decide on the preferred option(s), and coordinate access to selected service(s). Veterans in the PNP had follow-up appointments with their Pain Navigator at 6 and 12 weeks post-initial encounter. Additional details for the PNP and metrics for its usage have been previously described. [16]

The 4-item short form PROMIS® Physical Function and Pain Interference were the primary outcome measures for the AIM-Back study. A selected set of additional outcomes were collected to further inform the effectiveness of the program.[11]

### Patient and Public Involvement

The SCP and PNP were reviewed and modified in the planning stages of this trial based on input from our Veteran Research Engagement Panel (VetREP) as well as other partners including administrators, and clinical providers. Details on this process have been described in a prior publication.[17]

### Participating Clinics Eligibility, Recruitment, and Randomization

Veterans Affairs clinics were recruited to participate in AIM-Back in two waves from February 2020-September 2020 and from March 2021-December 2021. Primary Care clinics were eligible to participate in AIM-Back if they could provide clinical personnel to deliver either of the treatment interventions and had seen between 800 and 5000 unique patients diagnosed with LBP in the previous year, with two exceptions decided by study team as allowable during the site recruitment phase. The first exception was for a rural serving clinic with LBP visits slightly below 800 (n=733) to help diversify the location of our participating sites. The other exception was made for a clinic that had >5000 visits because we identified specific patient aligned care teams (PACT) within their primary care clinic to participate and maintain an appropriate eligible participant volume. The number of primary care personnel trained to refer to AIM-Back during the trial ranged from 4-69 per enrolled clinic.

The AIM-Back trial received site participation agreements from and randomized 19 VA Primary Care clinics to one of the two pathways. Two clinics withdrew in the early stages of rolling out the AIM-Back program and data from these clinics are excluded from all analyses. The 17 clinics resided in ten different states: West Virginia, South Carolina, Nevada, Missouri, Texas, Pennsylvania, North Carolina, Kansas, Ohio, and Kentucky. Participating clinics were randomized into two blocks using a blocked covariate constrained randomization [11] and, after randomization, each clinic underwent training on how to implement their assigned pathway. Block 1 consisted of ten clinics (five in each pathway) that were randomized in September 2020, with the first clinic actively referring Veterans in February 2021. Block 2 randomization was completed in December 2021 and included five PNP clinics and four SCP clinics, with the first clinic referring Veterans in March 2022. The 2 clinics that withdrew (described above), one from each pathway, were both from Block 2.

### Cohort and Sample Definition

To be eligible for AIM-Back, Veterans were referred to the randomized pathway at their clinic by trained primary care providers across 17 clinics (9 PNP; 8 SCP) between February 1, 2021 and January 31, 2024 (Referral Cohort). Veterans from the Referral cohort (n=2767; 1481 PNP; 1286 SCP) that participated in the study included: 1) an Electronic Health Record (EHR) sample of Veterans (n=1817; 1006 PNP, 811 SCP) enrolled in the program (i.e., attended initial AIM-Back visit with no consent required), approximately 66% of the Referral cohort, and 2) a Survey sample of Veterans (n=996; 480 PNP; 516 SCP) that agreed at the AIM-Back referral to be contacted about the research survey study were consented and completed baseline surveys (see Figure 1). Approximately 80% of the Referral cohort agreed to be contacted for the survey study (n=2183), 45.6% of whom were included in the survey sample There were n=799 Veterans in the EHR sample (44%) that were also in the Survey sample (see Figure 1); we planned for approximately 50% of the EHR sample to be in the Survey sample in the design of the study.

**Figure 1.**
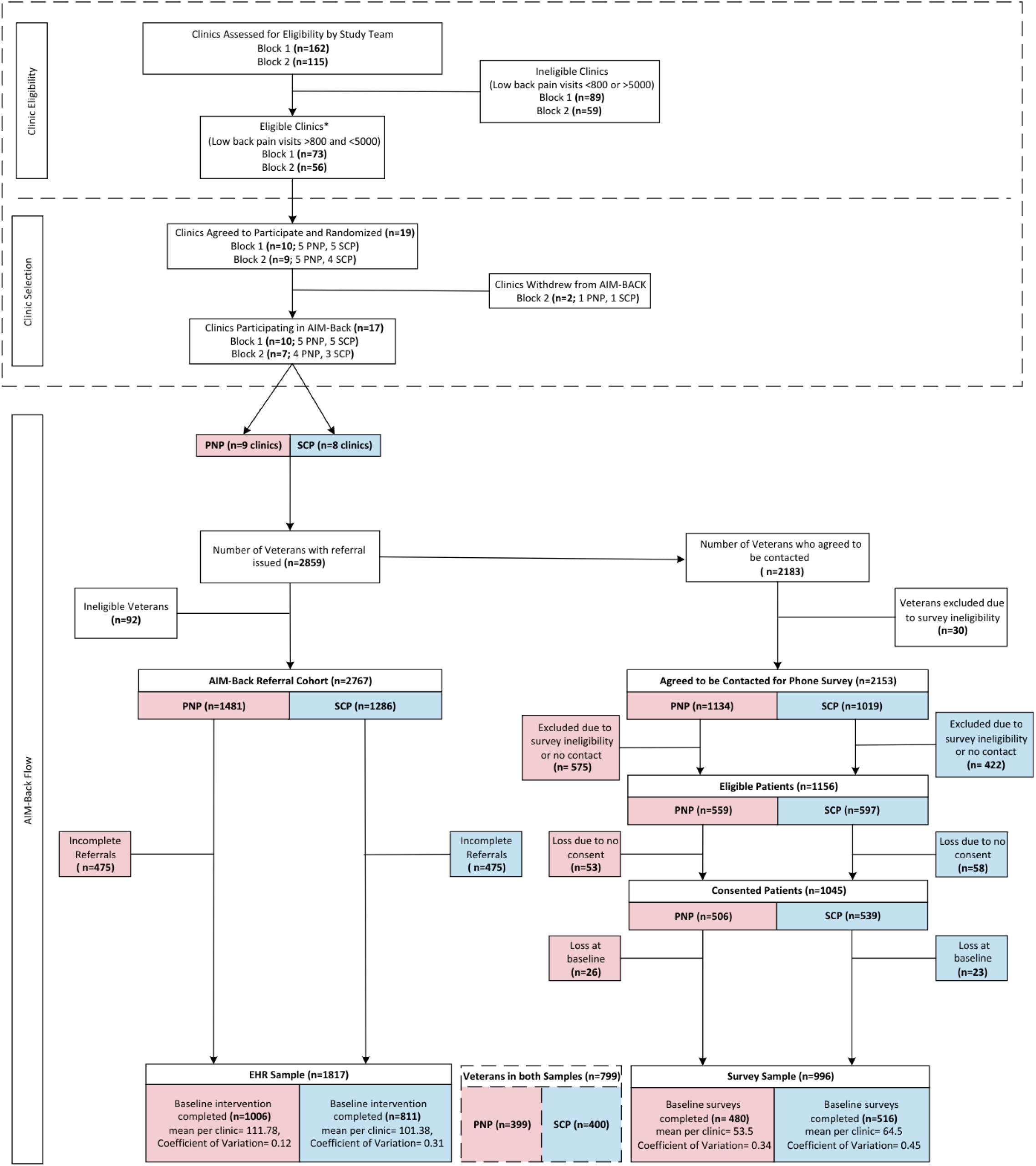
AIM-Back Baseline CONSORT

Each clinic had a targeted enrollment of 105 Veterans for the EHR sample with a minimum enrollment target of 65 and a maximum of 130 Veterans to minimize cluster imbalance; for the Survey sample the target enrollment was n=54 per clinic.[18] Trial enrollment concluded in January 2024.

Providers at participating clinics were trained to refer Veterans to AIM-Back who had low back pain, received primary care at a randomized clinic, had a valid telephone number, were not receiving or had not been referred to hospice or palliative care, and were informed that AIM-Back was best suited for Veterans for whom conservative care was appropriate. Individual consent was not obtained for Veterans in the EHR sample to participate because each clinic had agreed to deliver the guideline-concordant AIM-Back program as part of their usual clinical care; therefore, the care delivery was not considered research. To be eligible to be contacted about participating in the Survey sample from research staff, Veterans referred to AIM-Back had to agree, at the referral, to be contacted for the research study that collected more granular data in surveys with additional longer follow-up time points. Further eligibility criteria for the survey sample included not being in institutional care (nursing home or hospital), no cognitive impairment, dementia, or lack of decision-making capacity, no serious mental illness (diagnosis of schizophrenia, bipolar disorder, psychiatric hospitalization) in the previous year or no current high-risk suicide flag in their medical record, and available access to, with ability to communicate on, a telephone. After confirming some of the additional eligibility criteria via an EHR chart review, the study team attempted to call to further screen for any remaining eligibility and, if eligible, obtained consent and administered the baseline survey. Further details on survey enrollment are detailed in the protocol paper.[11]

### Methods of Data Collection

The AIM-Back trial had two data sources: 1) data entered through the Computerized Patient Record System (CPRS), the Veterans Health Administration Health Care System (VHAHCS) EHR, and extracted from the Corporate Data Warehouse (CDW) and 2) data collected via telephone by AIM-Back research staff.

Primary care clinicians referred patients to the pathway randomized to their clinic through the EHR. Once an AIM-Back referral was issued, clinics’ usual care protocols were followed to schedule the baseline intervention visit by an AIM-Back trained physical therapist or pain navigator (depending on the pathway to which the clinic was randomized). Demographic information (age, race, ethnicity, and sex), opioid use based on a prescription fill in the year prior to referral, PTSD diagnoses in year prior to referral, and Care Assessment Needs (CAN) scores prior to referral date (closest)[19] were extracted from CDW for the Referral cohort (see list of drugs for opioid use and ICD10 codes for PTSD diagnosis in Supplemental Materials). CAN score is a comorbidity measure and is the estimated risk for hospital admission or death within a year calculated weekly for all eligible Veterans. The score is the risk percentile and ranges from 0 (lowest risk) to 99 (highest risk). During the initial evaluation, the provider used a clinical note template specific to the clinic’s assigned AIM-Back pathway to collect data that included the 2-item CDC high impact pain questionnaire [15], the 2-item NIH Task Force on Research Standards for Chronic Low Back Pain [20], the 4-item PROMIS Pain Interference [21], the 4-item PROMIS Physical Function [22], the 4-item PROMIS Sleep Disturbance[23], and other relevant care notes. The data from these EHR templates are stored as structured text fields and uploaded nightly to CDW.

For the survey sample, patient demographics (baseline only), PROMIS pain interference, PROMIS physical function, PROMIS sleep disturbance, CDC and NIH definitions of HICP, Catastrophizing, PEG pain intensity, Self-efficacy, Depression (PHQ-2, AUDIT-C, Quality of life (EUROQOL), and Nonpharmacological and Self-Care Approaches (NSCAP)[24,25] measures were collected. Data were collected electronically into REDCap [26] at baseline, 3, 6, and 12 months.

In addition, to evaluate representativeness of the Referral Cohort for each clinic, we identified all primary care LBP visits based on ICD-10 codes (see list in Supplemental Materials) during the study active periods at each clinic and extracted demographics (age, race, ethnicity, and sex) for the Veterans with the LBP visits.

## Findings to Date

There are no longitudinal findings reported to date from the AIM-Back trial. Prior reports include a summary of our partner engagement process to refine the pathways included in the trial [17] and descriptive analyses of SCP and PNP use.[15,16] Trial enrollment has been completed and reporting of longitudinal findings, including comparative effectiveness of care pathways and treatment heterogeneity analyses, will follow baseline characteristics reporting.

Findings from this Cohort Profile paper are meant to provide foundational data to support later analyses from the AIM-Back trial. The descriptive data presented in this paper provide a comprehensive summary of the measures we collected via EHR and survey. Additionally, this Cohort Profile allows us to report in detail on key measures collected via survey that will be used in secondary analyses.

Age, sex, ethnicity, and race demographics are presented in Table 1 alongside rates of HICP, PTSD, and opioid use for the Referral cohort, EHR and Survey samples overall, and by pathway. The mean ages for the Referral Cohort, Survey sample and the EHR were similar (51-53 years old). The Referral Cohort was 11.8% female, 28.5% Black/African American, 10.5% had documented opioid use in the year prior, and 23.7% had a documented PTSD ICD-10 code. The EHR sample was 12.1% female, 29.3% Black/African American, 9.9% had documented opioid use in the year prior, and 23.4% had a documented PTSD ICD-10 code. The Survey sample was 10.9% female and 25.0% Black/African American, 9.1% had documented opioid use in the year prior, and 24.7% had a documented PTSD ICD-10 code. In the EHR sample, opioid use was slightly higher among those in the PNP (10.8%) than those in the SCP (8.6%), and there was variability across clinics with a minimum opioid use of 2.4% and a maximum opioid use of 15.3% assessed in the year prior to referral (Coefficient of Variation (CV)=0.40).

**Table 1.**
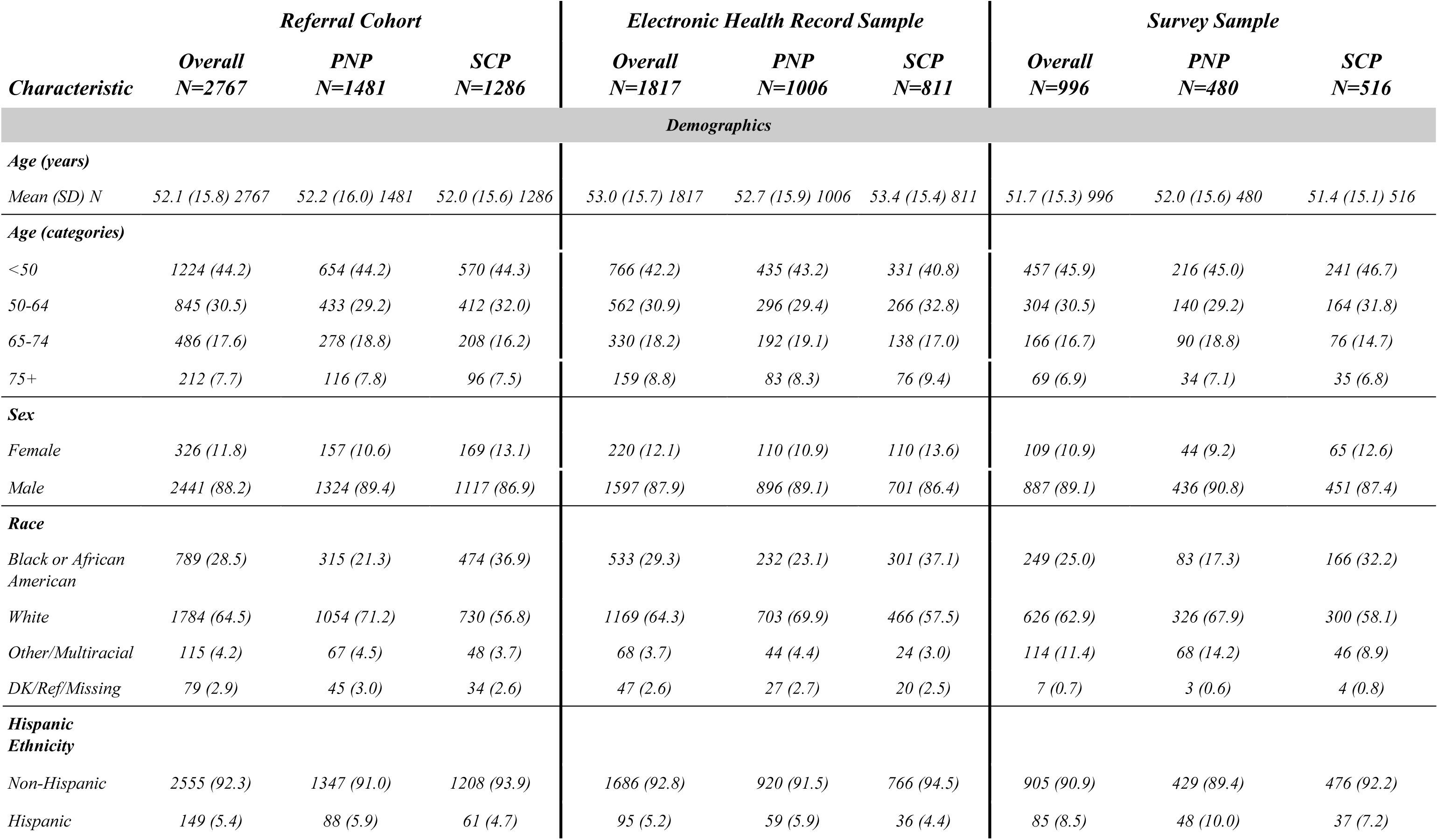
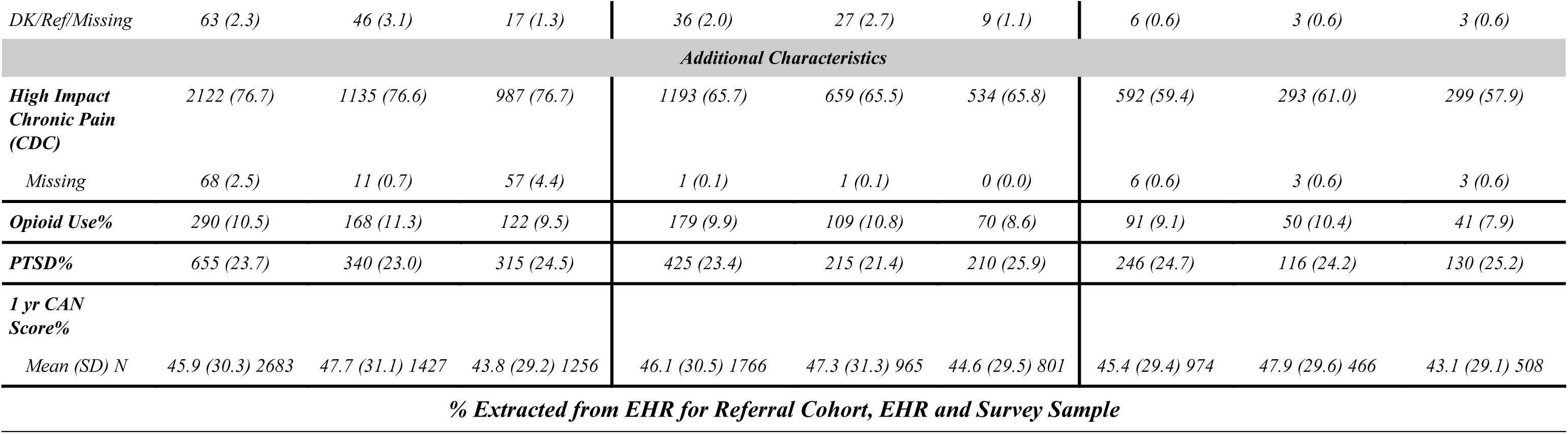
Baseline Characteristics in the Referral Cohort and the EHR and Survey Samples. *N (%) unless specified otherwise

| *Characteristic* | *Referral Cohort* |  |  | *Electronic Health Record Sample* |  |  | *Survey Sample* |  |  |
| --- | --- | --- | --- | --- | --- | --- | --- | --- | --- |
|  | *Overall*<br>*N=2767* | *PNP*<br>*N=1481* | *SCP*<br>*N=1286* | *Overall*<br>*N=1817* | *PNP*<br>*N=1006* | *SCP*<br>*N=811* | *Overall*<br>*N=996* | *PNP*<br>*N=480* | *SCP*<br>*N=516* |
| *Demographics* |  |  |  |  |  |  |  |  |  |
| *Age (years)* |  |  |  |  |  |  |  |  |  |
| *Mean (SD) N* | 52.1 (15.8) 2767 | 52.2 (16.0) 1481 | 52.0 (15.6) 1286 | 53.0 (15.7) 1817 | 52.7 (15.9) 1006 | 53.4 (15.4) 811 | 51.7 (15.3) 996 | 52.0 (15.6) 480 | 51.4 (15.1) 516 |
| *Age (categories)* |  |  |  |  |  |  |  |  |  |
| <50 | 1224 (44.2) | 654 (44.2) | 570 (44.3) | 766 (42.2) | 435 (43.2) | 331 (40.8) | 457 (45.9) | 216 (45.0) | 241 (46.7) |
| 50-64 | 845 (30.5) | 433 (29.2) | 412 (32.0) | 562 (30.9) | 296 (29.4) | 266 (32.8) | 304 (30.5) | 140 (29.2) | 164 (31.8) |
| 65-74 | 486 (17.6) | 278 (18.8) | 208 (16.2) | 330 (18.2) | 192 (19.1) | 138 (17.0) | 166 (16.7) | 90 (18.8) | 76 (14.7) |
| 75+ | 212 (7.7) | 116 (7.8) | 96 (7.5) | 159 (8.8) | 83 (8.3) | 76 (9.4) | 69 (6.9) | 34 (7.1) | 35 (6.8) |
| *Sex* |  |  |  |  |  |  |  |  |  |
| *Female* | 326 (11.8) | 157 (10.6) | 169 (13.1) | 220 (12.1) | 110 (10.9) | 110 (13.6) | 109 (10.9) | 44 (9.2) | 65 (12.6) |
| *Male* | 2441 (88.2) | 1324 (89.4) | 1117 (86.9) | 1597 (87.9) | 896 (89.1) | 701 (86.4) | 887 (89.1) | 436 (90.8) | 451 (87.4) |
| *Race* |  |  |  |  |  |  |  |  |  |
| *Black or African American* | 789 (28.5) | 315 (21.3) | 474 (36.9) | 533 (29.3) | 232 (23.1) | 301 (37.1) | 249 (25.0) | 83 (17.3) | 166 (32.2) |
| *White* | 1784 (64.5) | 1054 (71.2) | 730 (56.8) | 1169 (64.3) | 703 (69.9) | 466 (57.5) | 626 (62.9) | 326 (67.9) | 300 (58.1) |
| *Other/Multiracial* | 115 (4.2) | 67 (4.5) | 48 (3.7) | 68 (3.7) | 44 (4.4) | 24 (3.0) | 114 (11.4) | 68 (14.2) | 46 (8.9) |
| *DK/Ref/Missing* | 79 (2.9) | 45 (3.0) | 34 (2.6) | 47 (2.6) | 27 (2.7) | 20 (2.5) | 7 (0.7) | 3 (0.6) | 4 (0.8) |
| *Hispanic Ethnicity* |  |  |  |  |  |  |  |  |  |
| *Non-Hispanic* | 2555 (92.3) | 1347 (91.0) | 1208 (93.9) | 1686 (92.8) | 920 (91.5) | 766 (94.5) | 905 (90.9) | 429 (89.4) | 476 (92.2) |
| *Hispanic* | 149 (5.4) | 88 (5.9) | 61 (4.7) | 95 (5.2) | 59 (5.9) | 36 (4.4) | 85 (8.5) | 48 (10.0) | 37 (7.2) |
| DK/Ref/Missing | 63 (2.3) | 46 (3.1) | 17 (1.3) | 36 (2.0) | 27 (2.7) | 9 (1.1) | 6 (0.6) | 3 (0.6) | 3 (0.6) |
| Additional Characteristics |  |  |  |  |  |  |  |  |  |
| High Impact Chronic Pain (CDC) | 2122 (76.7) | 1135 (76.6) | 987 (76.7) | 1193 (65.7) | 659 (65.5) | 534 (65.8) | 592 (59.4) | 293 (61.0) | 299 (57.9) |
| Missing | 68 (2.5) | 11 (0.7) | 57 (4.4) | 1 (0.1) | 1 (0.1) | 0 (0.0) | 6 (0.6) | 3 (0.6) | 3 (0.6) |
| Opioid Use% | 290 (10.5) | 168 (11.3) | 122 (9.5) | 179 (9.9) | 109 (10.8) | 70 (8.6) | 91 (9.1) | 50 (10.4) | 41 (7.9) |
| PTSD% | 655 (23.7) | 340 (23.0) | 315 (24.5) | 425 (23.4) | 215 (21.4) | 210 (25.9) | 246 (24.7) | 116 (24.2) | 130 (25.2) |
| 1 yr CAN Score% |  |  |  |  |  |  |  |  |  |
| Mean (SD) N | 45.9 (30.3) 2683 | 47.7 (31.1) 1427 | 43.8 (29.2) 1256 | 46.1 (30.5) 1766 | 47.3 (31.3) 965 | 44.6 (29.5) 801 | 45.4 (29.4) 974 | 47.9 (29.6) 466 | 43.1 (29.1) 508 |
| % Extracted from EHR for Referral Cohort, EHR and Survey Sample |  |  |  |  |  |  |  |  |  |

The Survey sample has a higher percentage of Hispanic/Latino participants and lower rates of HICP prevalence compared to the Referral Cohort and EHR sample. The Referral cohort had the highest rates of HICP (76.7%) compared to the EHR Sample (65.7%) and the Survey sample (59.4%). There was variation across the 17 clinics in HICP with a Referral cohort range of 49.1-94.6%, an EHR sample range of 56.3-78.5%, and a Survey sample range of 43.8-74.0%. Additional findings related to HICP can be found in Table 1.

Table 2 contains the mean and standard deviation for the PROMIS T-scores by sample and pathway. The EHR sample had PROMIS Pain Interference and Physical Function mean T-scores of 63.2 and 37.1. Similarly, Veterans in the Survey sample had a mean Pain Interference T-score of 63.1 and a mean Physical Function T-score of 38.1. These scores are indicative of a moderate level of impairment for our co-primary outcome measures. Baseline mean PROMIS scores were similar across pathways for both the EHR and Survey sample.

**Table 2.**
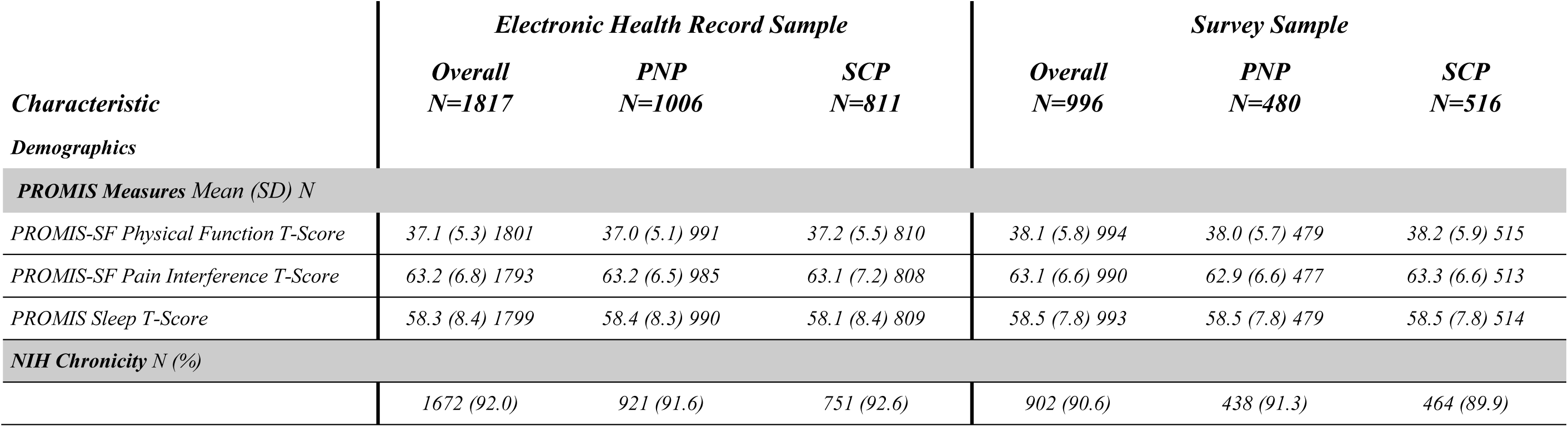
PROMIS Measures and NIH Chronicity.

Additional characteristics are available in Table 3 for those in the Survey sample to provide additional details about the Veterans participating in AIM-Back.

**Table 3.**
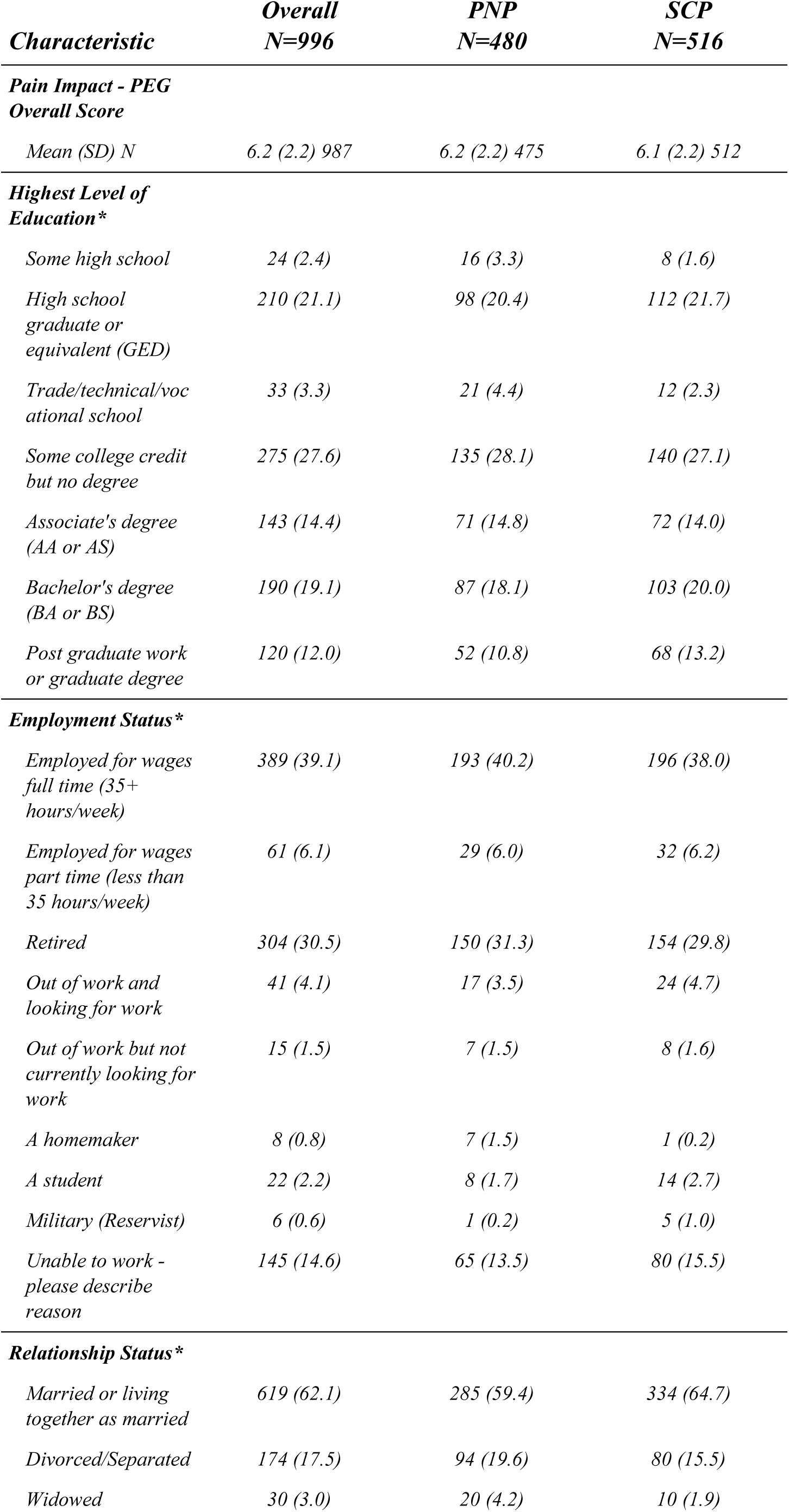
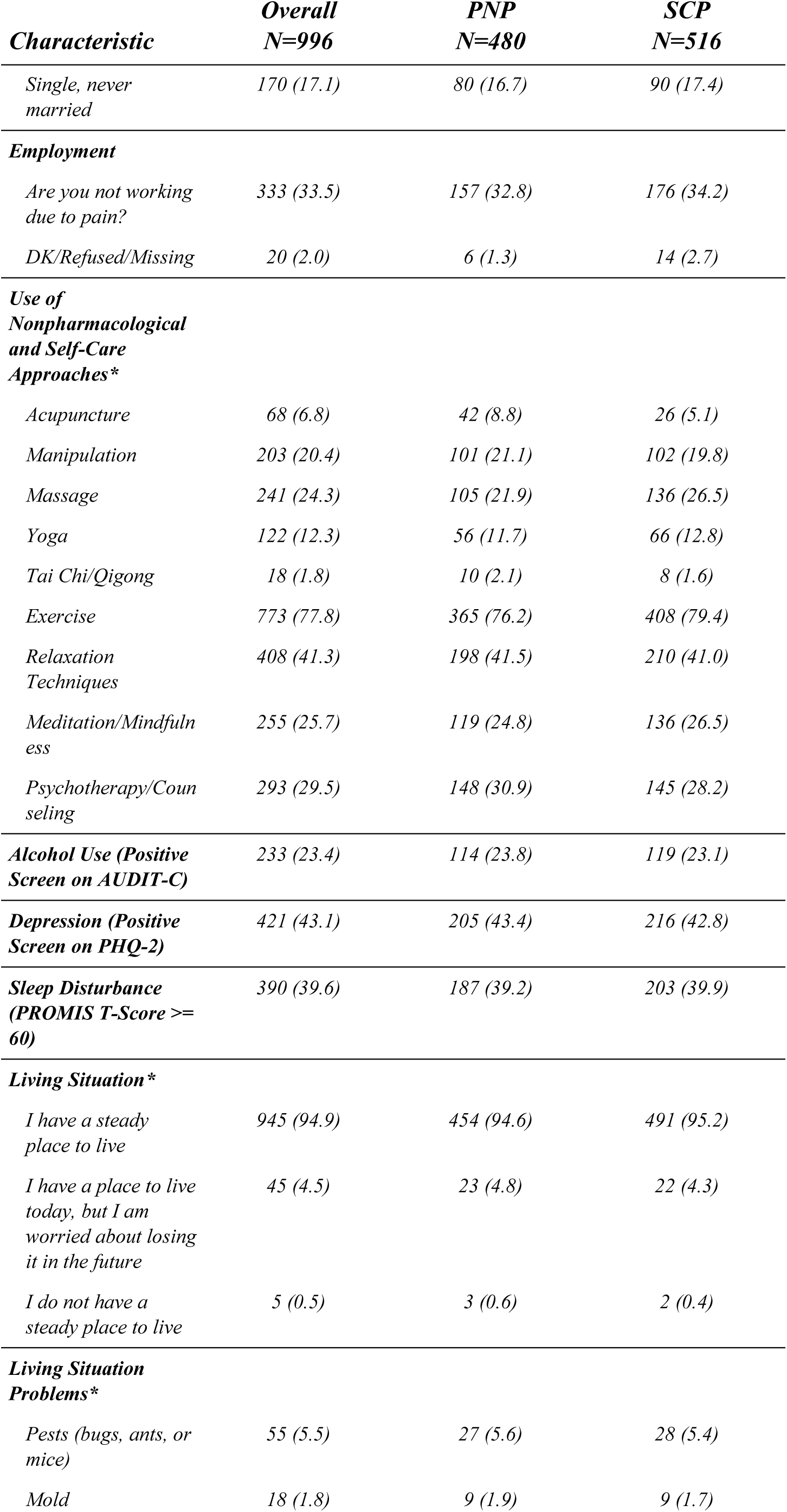
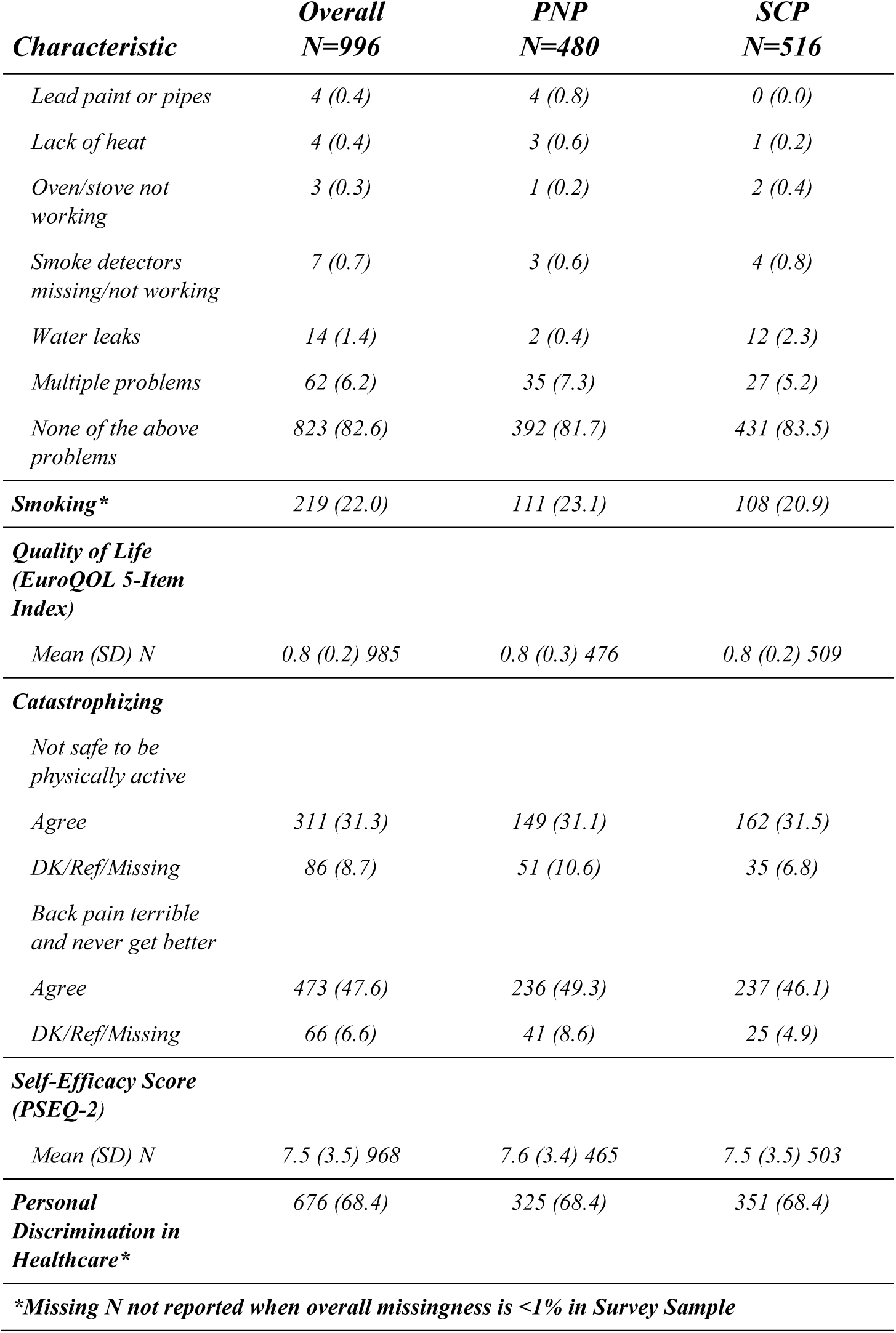
Additional Survey Sample Characteristics Survey Sample.

**Table 4.**
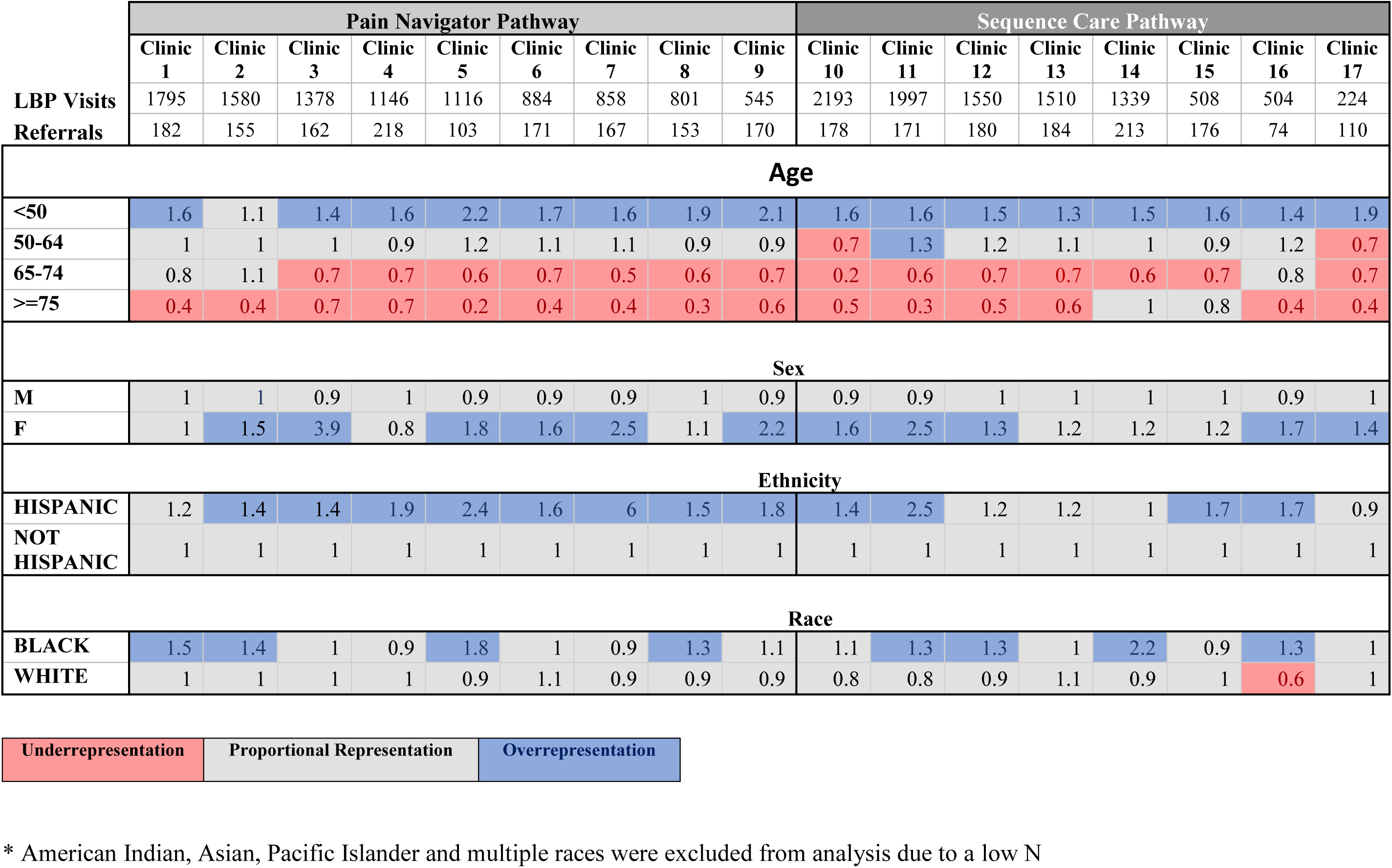
Participation to Prevalence Ratio.

To evaluate the representativeness of the Referral cohort, we calculated the participation to prevalence ratio (PPR) for age, race, sex, and ethnicity. PPR is a ratio of those within a specific demographic who participated relative to those of that same demographic who were eligible to participate from the local clinical environment.[27] For AIM-Back, the eligible participants were all Veterans with a documented LBP visit at a randomized primary care clinic during the active trial period. A PPR value between 0.8 and 1.2 indicates a likely representative sample, while values below 0.8 suggest potential for underrepresentation and values above 1.2 indicate potential for overrepresentation.[27] As shown in Table 3, Veterans aged 50 or younger were consistently overrepresented in referrals to AIM-Back by providers across the 17 clinics, referrals of female Veterans were often overrepresented, and referrals occasionally overrepresented Black/African American Veterans. Conversely, referrals to AIM-back during the trial consistently underrepresented Veterans aged 65+ across clinics.

HICP and opioid usage are reported as factors that will be used for planned subgroup analyses. The elevated presence of HICP provides an initial indication that the AIM-Back samples consist of Veterans with high levels of interference from their LBP, which will serve as a highly relevant characteristic in interpreting findings from the primary analyses. The prevalence of HICP and opioid usage suggests that we will have enough participants to proceed with the planned subgroup analyses.

Generalizability is a key issue in interpreting clinical trial results yet is often hard to assess empirically. In this Cohort Profile paper, we used the PPR as an indicator to assess representativeness of those referred to AIM-Back by trained primary care providers to the LBP population at each clinic.

Overall, we identified that Veterans referred to the AIM-Back program were representative based on sex and race. However, we also had evidence across multiple enrolling clinics that older Veterans (65+ years) were underrepresented for referral to AIM-Back. This finding was unexpected and could be due to the way we implemented the programs (e.g., without an emphasis on including older Veterans) or the perception of referring providers on the treatment delivered (e.g., the care pathways were not well suited for older Veterans). At this point, the reasons for this finding are speculative but will be important to know for those interested in Veteran populations for whom these trial findings will be most relevant and for any future implementation efforts aligned with these care pathways.

### Strengths and Limitations

The findings reported here indicate that the AIM-Back trial has several strengths, including a) having data from the Referral Cohort to understand provider and patient engagement with these pathways; b) the enrollment goal was met, which allows for adequate power of all planned analyses; c) the use of both EHR and survey samples provide for a comprehensive assessment, allowing for future robust evaluation and sensitivity analyses of trial findings; and d) the integration of AIM-Back features within existing clinical settings demonstrates the potential for these care pathways to be implemented more broadly in Veteran Health Administration clinics and in other healthcare environments outside the Veteran Health Administration. Additionally, since cluster randomization can result in imbalances, it is encouraging to see balance in our primary and secondary outcome measures at baseline across care pathways.[18]

Some of the limitations of the trial include recruitment during the COVID-19 pandemic that affected practice patterns for clinics participating during shutdowns compared to care delivered pre- and post-shutdown. Leveraging the EHR as a data collection tool presented challenges related to consistent documentation which may impact our analysis, such as the PPR, that is reliant on ICD-10 code documentation for LBP visits which is known to be inconsistently used across healthcare systems and personnel. [28] Lack of appropriate diagnostic documentation may miss detection of Veterans eligible to receive AIM-Back. An important consideration for future research and implementation efforts is to further explore participation among older Veterans to overcome the current limitations of the trial’s generalizability due to the lower referral of this subgroup to the AIM-Back program.

### Collaboration

Deidentified data will be made available upon request and permission for use granted consistent with VA policy via inquiry to.

## Data Availability

All data produced in the present study are available upon reasonable request to the authors, after the primary outcomes are reported in a peer-reviewed manuscript

## Acknowledgments

This work is supported through cooperative agreement UH3AT009790 from the NIH, National Center for Complimentary and Integrative Health (NCCIH). The content is solely the responsibility of the authors and does not necessarily represent the official views of the National Institutes of Health the U.S Department of Veterans Affairs or the United States Government

This manuscript is a product of the Pain Management Collaboratory. For more information about the Collaboratory, visit https://painmanagementcollaboratory.org/.”

This material is the result of work supported with resources and the use of facilities at the Durham VA Health Care System in Durham North Carolina.

## Supplemental Materials

### Low Back Pain ICD-10 codes

|  |  |
| --- | --- |
| M43.05 | Spondylolysis, thoracolumbar region |
| M43.06 | Spondylolysis, lumbar region |
| M43.07 | Spondylolysis, lumbosacral region |
| M43.15 | Spondylolisthesis, thoracolumbar region |
| M43.16 | Spondylolisthesis, lumbar region |
| M43.17 | Spondylolisthesis, lumbosacral region |
| M43.27 | Fusion of spine, lumbosacral region |
| M43.28 | Fusion of spine, sacral and sacrococcygeal region |
| M46.47 | Discitis, unspecified, lumbosacral region |
| M47.15 | Other spondylosis with myelopathy, thoracolumbar region |
| M47.16 | Other spondylosis with myelopathy, lumbar region |
| M47.25 | Other spondylosis with radiculopathy, thoracolumbar region |
| M47.26 | Other spondylosis with radiculopathy, lumbar region |
| M47.27 | Other spondylosis with radiculopathy, lumbosacral region |
| M47.815 | Spondylosis without myelopathy or radiculopathy, thoracolumbar region |
| M47.816 | Spondylosis without myelopathy or radiculopathy, lumbar region |
| M47.817 | Spondylosis without myelopathy or radiculopathy, lumbosacral region |
| M47.895 | Other spondylosis, thoracolumbar region |
| M47.896 | Other spondylosis, lumbar region |
| M47.897 | Other spondylosis, lumbosacral region |
| M48.05 | Spinal stenosis, thoracolumbar region |
| M48.06 | Spinal Stenosis, Lumbar Region |
| M48.07 | Spinal stenosis, lumbosacral region |
| M48.20 | Kissing spine, site unspecified |
| M48.8X5 | Other specified spondylopathies, thoracolumbar region |
| M51.05 | Intervertebral disc disorders with myelopathy, thoracolumbar region |
| M51.06 | Intervertebral disc disorders with myelopathy, lumbar region |
| M51.15 | Intervertebral disc disorders with radiculopathy, thoracolumbar region |
| M51.16 | Intervertebral disc disorders with radiculopathy, lumbar region |
| M51.17 | Intervertebral disc disorders with radiculopathy, lumbosacral region |
| M51.25 | Other intervertebral disc displacement, thoracolumbar region |
| M51.26 | Other intervertebral disc displacement, lumbar region |
| M51.27 | Other intervertebral disc displacement, lumbosacral region |
| M51.35 | Other intervertebral disc degeneration, thoracolumbar region |
| M51.36 | Other intervertebral disc degeneration, lumbar region |
| M51.37 | Other intervertebral disc degeneration, lumbosacral region |
| M51.85 | Other intervertebral disc disorders, thoracolumbar region |
| M51.86 | Other intervertebral disc disorders, lumbar region |
| M51.87 | Other intervertebral disc disorders, lumbosacral region |
| M51.9 | Unspecified thoracic, thoracolumbar and lumbosacral intervertebral disc disorder |
| M53.2X7 | Spinal instabilities, lumbosacral region |
| M53.3 | Sacrococcygeal disorders, not elsewhere classified |
| M54.15 | Radiculopathy, thoracolumbar region |
| M54.16 | Radiculopathy, lumbar region |
| M54.17 | Radiculopathy, lumbosacral region |
| M54.30 | Sciatica, unspecified side |
| M54.31 | Sciatica, right side |
| M54.32 | Sciatica, left side |
| M54.40 | Lumbago with sciatica, unspecified side |
| M54.41 | Lumbago with sciatica, right side |
| M54.42 | Lumbago with sciatica, left side |
| M54.5 | Low back pain |
| M54.50 | Low back pain, unspecified |
| M96.1 | Postlaminectomy syndrome, not elsewhere classified |
| M99.03 | Segmental and somatic dysfunction of lumbar region |
| M99.04 | Segmental and somatic dysfunction of sacral region |
| M99.13 | Subluxation complex (vertebral) of lumbar region |
| Q76.2 | Congenital spondylolisthesis |
| S33.5XXA | Sprain of ligaments of lumbar spine, initial encounter |
| S33.6XXA | Sprain of sacroiliac joint, initial encounter |
| S33.8XXA | Sprain of other parts of lumbar spine and pelvis, initial encounter |
| S33.9XXA | Sprain of unspecified parts of lumbar spine and pelvis, initial encounter |

### Post-Traumatic Stress Disorder ICD-10 codes

|  |  |
| --- | --- |
| F43.10 | Post-traumatic stress disorder, unspecified |
| F43.11 | Post-traumatic stress disorder, acute |
| F43.12 | Post-traumatic stress disorder, chronic |

### Prescription drugs used to define Opioid Use

|  |  |
| --- | --- |
| Butorphanol | Methadone |
| Codeine | Nalbuphine |
| Dihydrocodeine | Oxycodone |
| Fentanyl | Morphine |
| Hydrocodone | Oxymorphone |
| Hydromorphone | Opium |
| Levorphanol | Pentazocine |
| Meperidine | Propoxyphene |
| Tapentadol | Tramadol |

